# Plasma biomarkers associated with survival and thrombosis in hospitalized COVID-19 patients

**DOI:** 10.1101/2021.11.10.21266185

**Authors:** David Cabrera-Garcia, Andrea Miltiades, Peter Yim, Samantha Parsons, Katerina Elisman, Mohammad Taghi Mansouri, Gebhard Wagener, Neil L. Harrison

**Affiliations:** Department of Anesthesiology, Columbia University Irving Medical Center, 630 West 168th Street, New York, NY 10032; Department of Molecular Pharmacology and Therapeutics, Columbia University Irving Medical Center, 630 West 168th Street, New York, NY 10032

**Keywords:** Coagulopathy, fibrinolysis, PAI-1, vWF, ST2, COVID-19

## Abstract

Severe coronavirus disease-19 (COVID-19) has been associated with fibrin-mediated hypercoagulability and thromboembolic complications. To evaluate potential biomarkers of coagulopathy and disease severity in COVID-19, we measured plasma levels of eight biomarkers potentially associated with coagulation, fibrinolysis, and platelet function in 43 controls and 63 COVID-19 patients, including 47 patients admitted to the intensive care unit (ICU) and 16 non-ICU patients. COVID-19 patients showed significantly elevated levels of fibrinogen, tissue plasminogen activator (t-PA), and its inhibitor plasminogen activation inhibitor 1 (PAI-1), as well as ST2 (the receptor for interleukin 33) and von Willebrand factor (vWF) compared to the control group. We found that higher levels of t-PA, ST2, and vWF at the time of admission were associated with lower survival rates, and that thrombotic events were more frequent in patients with initial higher levels of vWF. These results support a predictive role of specific biomarkers such as t-PA and vWF in the pathophysiology of COVID-19. The data provide support for the case that hypercoagulability in COVID-19 is fibrin-mediated, but also highlights the important role that vWF may play in the genesis of thromboses in the pathophysiology of COVID-19. Interventions designed to enhance fibrinolysis and reduce platelet aggregation might prove to be useful adjuncts in the treatment of coagulopathy in a subset of COVID-19 patients.

## 1. Introduction

COVID-19, the disease associated with infection by the severe acute respiratory syndrome coronavirus 2 (SARS-CoV-2), can cause upper respiratory tract (URT) infections that may progress to bilateral pneumonia and acute respiratory distress syndrome (ARDS) [1]. Extra-pulmonary symptoms are also a characteristic of patients with severe COVID-19, and these include systemic inflammation and cytokine release [2], often combined with disorders of blood clotting and secondary manifestations of disease in the lung, kidney, brain, heart, and liver [3]. Increased incidence of thrombosis and thrombotic complications have been widely associated with COVID-19 [4,5]. Changes in conventional coagulation parameters such as international normalized ratio (INR), D-dimer, and platelet counts may be useful for the rapid detection of hypocoagulation in COVID-19 patients [6], but these parameters do not fully describe the complex biology associated with COVID-19 associated coagulopathy [7].

Our clinical observations of increased thromboembolic complications in COVID-19 led us to use rotational thromboelastometry (ROTEM) to study the mechanisms of hypercoagulability in severe COVID-19. We found that there was a significant increase of fibrin-mediated clot viscosity [8], but the exact mechanism of this fibrin-mediated hypercoagulability has remained elusive. Understanding of the pathogenesis of coagulopathy in COVID-19 is incomplete, and treatment with anticoagulants such as heparin in severe COVID-19 patients has been used with mixed results [9]. We hypothesized that coagulopathy associated with severe COVID-19 might be associated with deficits in fibrinolysis, as well as with alterations in the primary coagulation pathway and platelet factors. We designed a study to measure selected coagulation-related biomarkers in plasma from patients with mild and severe COVID-19, admitted between April 2020 and April 2021.

## 2. Patients and methods

### 2.1. Patients

We conducted a prospective cohort study that was approved by Columbia University IRB (protocol #AAAS0172). Sixty-three patients with COVID-19 and 43 controls without COVID-19 were enrolled. COVID-19 status was confirmed by standard reverse transcription-polymerase chain reaction (RT-PCR) test for SARS-CoV-2. 47 of the COVID-19 patient group had symptoms of critical illness (severe infection, respiratory distress, shock, and/or multiorgan dysfunction), requiring admission to the intensive care unit (ICU), while the remaining 16 COVID-19 patients (non-ICU) were admitted to a non-acute hospital bed. The control patient group consisted of healthy volunteers and admitted surgical patients with no clinical evidence of viral infection and a negative RT-PCR test within 5 days of sample collection. Informed consent was provided by the patients or the surrogates and blood was drawn. Blood of the same-day surgical patients was drawn prior to the beginning of the surgical procedure. Volunteers were not remunerated for participating in the study and health status for the volunteers was determined by self-report. The study was performed at Columbia University Irving Medical Center in New York, N.Y., and COVID-19 patients and controls were recruited in two phases during April-May 2020, and between November 2020 and April 2021. Blood was drawn into EDTA-containing tubes, rapidly centrifuged and plasma separated before being stored at −80 °C until analysis. Samples from COVID-19 patients were collected between 5 and 15 days following admission to the ICU. No solvents or detergents were added to the plasma samples, which were handled in a biological safety cabinet under enhanced Bio Safety Level 2 (BSL-2) protocols.

### 2.2. Enzyme-linked immunosorbent assay (ELISA)

ELISA kits with colorimetric output were used to determine the plasma concentrations of PAI-1 (ab184863, Abcam), fibrinogen (ab241383, Abcam), plasminogen (ab108893, Abcam), tissue plasminogen activator (t-PA) (ab190812, Abcam), platelet factor 4 (PF4) (ab189573, Abcam), interleukin (IL) 1 receptor-like 1 (ST2) (ab254505, Abcam), von Willebrand factor (vWF) (ab223864, Abcam), and tryptase (EKU10581, Biomatik) according to the manufacturer’s instructions. The plates were read at 450 nm using an automated microplate reader (Biotek Epoch). For each subject, we report the mean value from at least two replicate determinations with a coefficient of variation (CV) lower than 20%. The plasma concentrations of each biomarker were determined using a standard curve, constructed and fit using a 4P logistic regression equation, and the assay values were corrected by the appropriate dilution factor in each case.

### 2.3. Statistics

All statistical analyses were performed using Prism 8 (GraphPad Software, Inc). Comparisons between the different COVID-19 and control groups were performed using Fisher’s test (categorical variables) or the non-parametric Mann-Whitney or Kruskal-Wallis tests (continuous variables). Blood levels of the biomarkers investigated are presented here as population medians, together with either the 95% confidence interval (CI) or the interquartile range (IQR). Additional detailed statistical information is presented in the respective Figure Legends.

## 3. Results

### 3.1. Demographics and clinical characteristics

Sixty-three patients (36 males and 27 females, median age 60, IQR 55 – 69 years) hospitalized with COVID-19 were included in this study. In addition, we recruited 43 controls (20 males and 23 females), median age 45 (IQR 30 – 61) years. The COVID-19 patients included 47 patients admitted to the ICU (severe COVID-19) and 16 hospitalized patients not requiring ICU admission.

The clinical characteristics of COVID-19 patients are listed in **Table 1**. No differences were found between the ICU and non-ICU COVID-19 patients in demographics or the prevalence of co-morbidities such as obesity or hypertension (**Table 1**). Medications given to each COVID-19 group are also reported in **Table 1**. At the time of blood collection, 46 of 47 (97.9%) ICU patients and 1 of 16 non-ICU patients were mechanically ventilated.

**Table 1.** Clinical characteristics of COVID-19 patients.

|  | Total |  | non-ICU |  | ICU |  | p-value |
| --- | --- | --- | --- | --- | --- | --- | --- |
|  | n | (%) | n | (%) | n | (%) |  |
| Demographics |  |  |  |  |  |  |  |
| Subjects | 63 | - | 16 | - | 47 | - | - |
| Female | 27 | (42.8) | 8 | (50.0) | 19 | (40.4) | 0.5665 |
| Age (median) | 60 | - | 59 | - | 61 | - | 0.9346 |
| Comorbidities |  |  |  |  |  |  |  |
| Obesity | 45 | (71.4) | 12 | (75.0) | 33 | (70.2) | >0.9999 |
| Hypertension | 37 | (58.7) | 7 | (43.7) | 30 | (63.8) | 0.2397 |
| Diabetes | 29 | (46.0) | 5 | (31.2) | 24 | (51.1) | 0.2468 |
| Chronic kidney disease | 9 | (14.2) | 1 | (6.2) | 8 | (17.0) | 0.4271 |
| Immunocompromised | 9 | (14.2) | 1 | (6.2) | 8 | (17.0) | 0.4271 |
| Heart diseases | 8 | (12.6) | 1 | (6.2) | 7 | (14.9) | 0.6673 |
| Lung diseases | 8 | (12.6) | 5 | (31.2) | 3 | (6.4) | **0.0277** |
| In-hospital medications |  |  |  |  |  |  |  |
| Steroids | 44 | (69.8) | 15 | (93.7) | 29 | (61.1) | **0.0247** |
| Broad spectrum antibiotics | 50 | (79.3) | 7 | (43.7) | 43 | (91.4) | **0.0002** |
| Azithromycin | 37 | (58.8) | 5 | (31.2) | 32 | (68.1) | **0.0173** |
| Hydroxychloroquine | 32 | (50.8) | 0 | (0.0) | 32 | (68.1) | **<0.0001** |
| Remdesivir | 20 | (31.7) | 10 | (62.5) | 10 | (21.3) | **0.0043** |
| IL-6 antibodies | 14 | (22.2) | 0 | (0.0) | 14 | (29.8) | **0.0133** |
| Anticoagulants | 16 | (25.4) | 4 | (25.0) | 12 | (25.5) | >0.9999 |
| Clinical outcomes |  |  |  |  |  |  |  |
| Ventilation | 46 | (73.0) | 0 | (0.0) | 46 | (97.9) | **<0.0001** |
| Acute kidney injury (AKI) | 40 | (63.9) | 5 | (31.2) | 35 | (74.5) | **0.0030** |
| AKI stage 1 | 12 | (19.0) | 5 | (31.2) | 7 | (14.9) | 0.1623 |
| AKI stage 2 | 4 | (6.9) | 0 | (0.0) | 4 | (8.5) | 0.5645 |
| AKI stage 3 | 24 | (38.1) | 0 | (0.0) | 24 | (51.1) | **0.0002** |
| Thrombotic events | 23 | (36.5) | 2 | (12.5) | 21 | (44.7) | **0.0355** |
| CRRT | 19 | (30.2) | 0 | (0.0) | 19 | (40.4) | **0.0014** |
| Deceased | 17 | (26.9) | 1 | (6.2) | 16 | (34.0) | **0.0479** |

Steroids received include dexamethasone, hydrocortisone, and/or methylprednisolone. IL-6 antibodies: Sarilumab (Kevzara) or tocilizumab (Actemra). Heart diseases include coronary artery disease and congestive heart failure. Ventilation status is reported at the time of blood collection. P-values are for non-ICU compared to ICU patients (Mann Whitney or Fisher's test). Values in bold are P < 0.05.

Thrombotic events, including stroke, were more common in ICU than in non-ICU patients (44.7% vs 12.5%, respectively). Similarly, a higher percentage of patients (74.5%) with acute kidney injury (AKI) was reported in ICU patients, and 19 ICU patients required continuous renal replacement therapy (CRRT). At the conclusion of this study (September 2021), 17 patients (27.0%) had died and the remainder of the patients, 46 (73.0 %), were discharged.

In **Table 2**, we report selected laboratory values for COVID-19 patients. As reported by others [10], levels of D-dimer were significantly elevated in ICU patients [IQR 2.0 to 11.5 mg/L]. D-dimer levels were higher in the group of patients admitted to the ICU than in non-ICU patients (P < 0.0001). Levels of bilirubin and serum creatinine were also higher in the ICU group.

**Table 2.** Laboratory results of COVID-19 patients.

|  | Reference values | All (n = 63) |  | non-ICU (n = 16) |  | ICU (n = 47) |  | p-value |
| --- | --- | --- | --- | --- | --- | --- | --- | --- |
|  |  | Median | (IQR) | Median | (IQR) | Median | (IQR) |  |
| **Coagulation markers** |  |  |  |  |  |  |  |  |
| INR | 0.8 – 1.2 | 1.2 | (1.1 – 1.3) | 1.2 | (1.0 – 1.3) | 1.2 | (1.2 – 1.3) | 0.1529 |
| Prothrombin time (s) | 20 – 35 | 34.3 | (30.8 – 54.6) | 31.1 | (29.1 – 33.9) | 36.3 | (31.2 – 61.2) | 0.0680 |
| D-dimer ($\mu\text{g/mL}$ FEU) | < 0.8 | 4.0 | (1.5 – 9.7) | 1.3 | (0.7 – 2.0) | 5.3 | (2.0 – 11.5) | **<0.0001** |
| **Blood count** |  |  |  |  |  |  |  |  |
| Platelets ($10^9/\text{L}$) | 150 – 350 | 271 | (189 – 350) | 276 | (193 – 347) | 271 | (184 – 350) | 0.5921 |
| White blood cells ($10^9/\text{L}$) | 4.5 – 11 | 11.5 | (7.4 – 15.8) | 9.4 | (4.9 – 14.3) | 12.1 | (8.0 – 16.9) | 0.1275 |
| Hemoglobin (g/dL) | 12 – 17 | 10.6 | (8.4 – 12.8) | 13.1 | (11.5 – 13.9) | 9.8 | (8.0 – 11.4) | **0.0001** |
| **Organ injury** |  |  |  |  |  |  |  |  |
| Bilirubin (mg/dL) | < 0.3 | 0.5 | (0.3 – 0.6) | 0.3 | (0.2 – 0.5) | 0.5 | (0.4 – 0.7) | **0.0091** |
| Serum creatinine (mg/dL) | 0.8 – 1.2 | 1.1 | (0.8 – 2.3) | 1 | (0.7 – 1.2) | 1.3 | (0.9 – 2.6) | **0.0212** |

Values are reported as medians (IQR). Laboratory tests were conducted within one day of the date of collection of blood samples for ELISA assays. Abbreviations: International normalized ratio (INR), fibrinogen equivalent units (FEU). Values in bold are $P < 0.05$ (Mann-Whitney test) for non-ICU vs ICU patients.

### 3.2. Coagulation and fibrinolysis markers in mild and severe COVID-19 patients

We measured the levels of fibrinogen, plasminogen, PAI-1, and t-PA in the plasma of the 63 COVID-19 patients and 43 controls (**Figure 1**) to assess the state of the coagulation and fibrinolytic systems. Fibrinogen levels in patients with non-ICU and ICU COVID-19 groups were significantly higher than in the control group (Control vs non-ICU: P < 0.0001, Control vs ICU: P < 0.0001) (**Figure 1a**). No difference was found in plasminogen levels between controls and COVID-19 patients (**Figure 1b**), but levels of t-PA were significantly higher in both COVID-19 groups compared to controls (Controls 4.0 ng/mL vs non-ICU COVID-19: 12.5 ng/mL; P < 0.0001, and vs ICU COVID-19: 22.3 ng/mL; P < 0.0001) **Figure 1c**). In addition, PAI-1 levels were elevated in ICU COVID-19 patients (**Figure 1d**).

**Figure 1.**
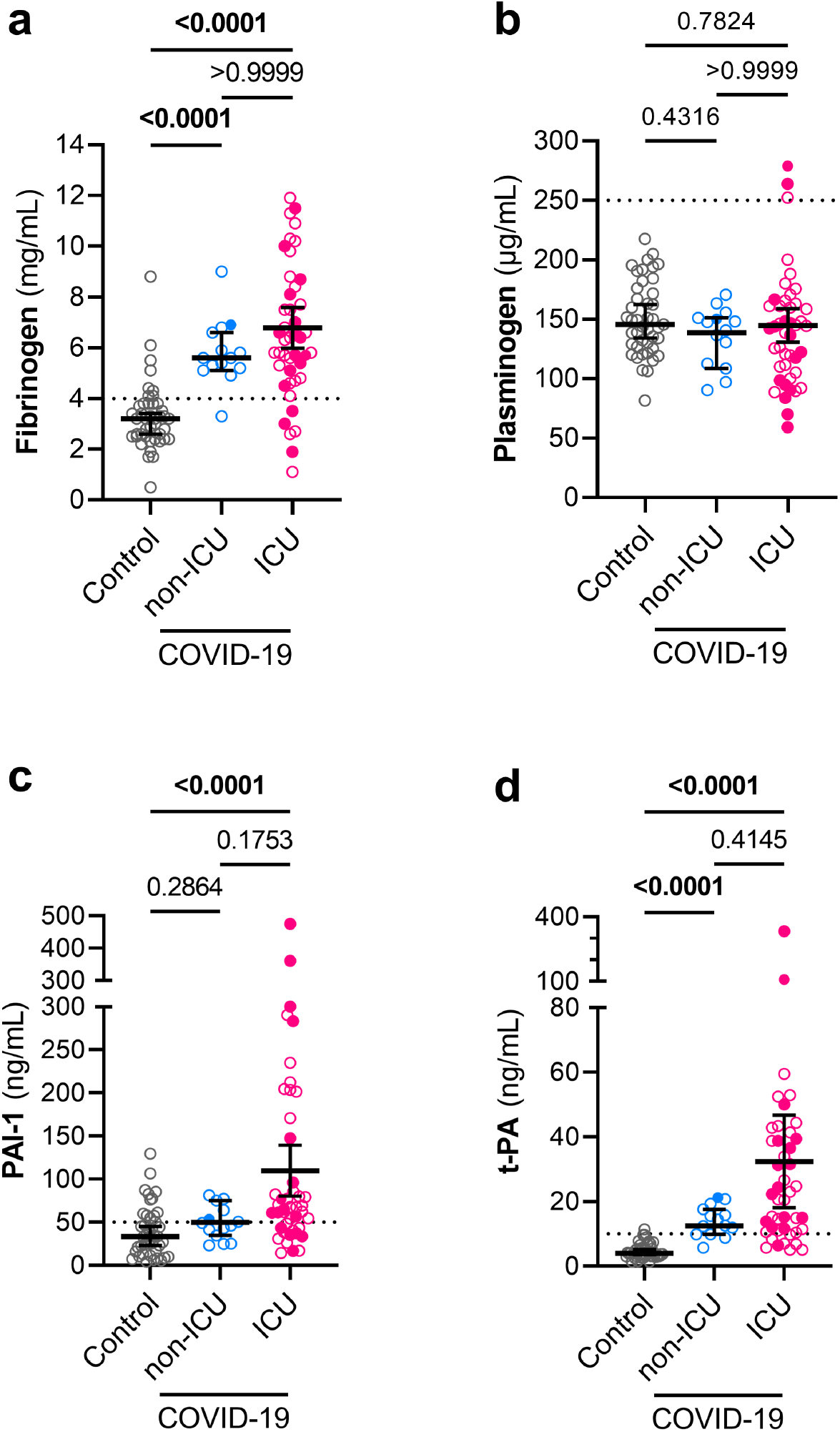
Plasma levels of fibrinogen, PAI-1, and t-PA are elevated in COVID-19 patients. Fibrinogen (**a**), plasminogen (**b**), PAI-1 (**c**), t-PA (**d**) were measured in plasma samples from controls (n = 43), non-ICU COVID-19 patients (n = 16) and ICU COVID-19 patients (n = 47). Graphs show individual values with the median and 95% confidence intervals. Deceased patients are indicated as filled symbols. Groups were compared using the Kruskal Wallis test and P-values are shown in each panel. Significant differences (P < 0.05) are in bold. The dotted lines represent the upper bound of the normal range for plasma levels of fibrinogen (4 mg/mL) (**a**), plasminogen (250 μg/mL) (**b**), PAI-1 (50 ng/mL) (**c**), and t-PA (10 ng/mL) (**d**).

### 3.3. Mast cell, T cell, and platelet-related factors in mild and severe COVID-19 patients

We also investigated the levels of ST2 (the receptor for interleukin-33) to assess the possible involvement of a variety of lymphocytes, including T cells [11], as well as the levels of tryptase as an index of recent mast cell degranulation [12]. Increased levels of ST2 were found in both groups of COVID-19 patients (P < 0.0001 for each group compared to controls), (**Figure 2a**). We observed somewhat lower levels of tryptase in the ICU COVID-19 group (**Figure 2b**). Finally, we measured the levels of vWF, a marker of endothelium damage and platelet adhesion [13], and the platelet activity factor PF4. Plasma levels of PF4 were similar in COVID-19 groups and controls (**Figure 2d**), but the levels of vWF were substantially elevated in both non-ICU and ICU COVID-19 groups (Controls: IQR 6.6 to 11.9 µg/mL vs non-ICU COVID-19: IQR 19.4 to 30 ng/mL; P = 0.0004, and vs ICU COVID-19: IQR 28.4 to 58.2 ng/mL; P < 0.0001) (**Figure 2c**).

**Figure 2.**
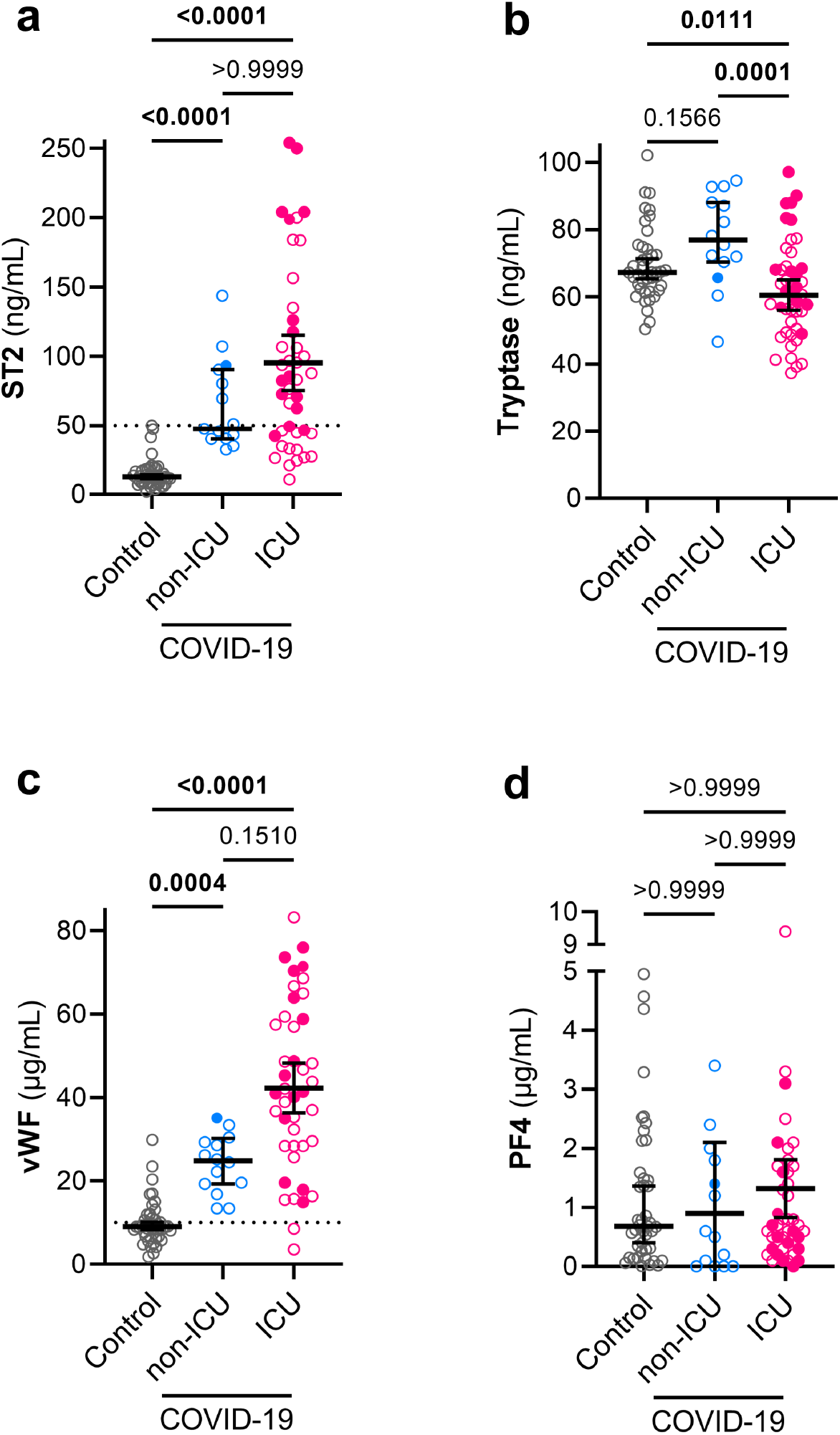
Plasma levels of ST2 and vWF are elevated in COVID-19 patients. ST2 (**a**), tryptase (**b**), vWF (**c**), and PF4 (**d**) were measured in plasma samples from control (n = 43), non-ICU COVID-19 (n = 16), and ICU COVID-19 patients (n = 47, n = 45 for **b-d**). Graphs show individual values with the median and 95% confidence intervals. Deceased patients are indicated with filled symbols. Groups were compared using the Kruskal-Wallis test and P-values are shown in each panel. Significant differences (P < 0.05) are in bold. The dotted lines represent the upper bound of the normal range for plasma levels of ST2 (50 ng/mL) (**a**) and vWF (10 μg/mL) (**d**).

We compared the plasma levels of these biomarkers with normal clinical ranges (dotted line in **Figure 1a, 1c, 1d, 2a, 2c**, respectively) and found that the majority of COVID-19 patients exhibited levels above the consensus normal range for fibrinogen (>4 mg/mL: 94% of non-ICU and 87% of ICU patients), t-PA (>10 ng/mL: 62% of non-ICU and 83% of ICU patients), PAI-1 (>50 ng/mL: 50% of non-ICU and 72% of ICU patients), ST2 (>50 ng/mL: 44% of non-ICU and 63% of ICU patients) and vWF (>10 µg/mL: 100% non-ICU and 92% of ICU patients).

### 3.4. High levels of t-PA, ST2, and vWF were associated with worse clinical outcomes in severe COVID-19

To investigate the predictive role of selected biomarkers, we asked whether there were any associations between the plasma levels of the biomarkers in COVID-19 patients at admission and two specific clinical outcomes (**Figure 3**). High levels of vWF were associated with thrombotic episodes (**Figure 3d**), which were more frequent among ICU patients (**Table 1, Figure 3**). Furthermore, higher levels of t-PA, ST2, and vWF at the time of admission were associated with lower likelihood of survival in COVID-19 patients (**Figure 3f-g**). In addition, we found that patients diagnosed with acute kidney injury (AKI) showed higher levels of t-PA and PAI-1 at the time of admission (**Figure S1**).

**Figure 3.**
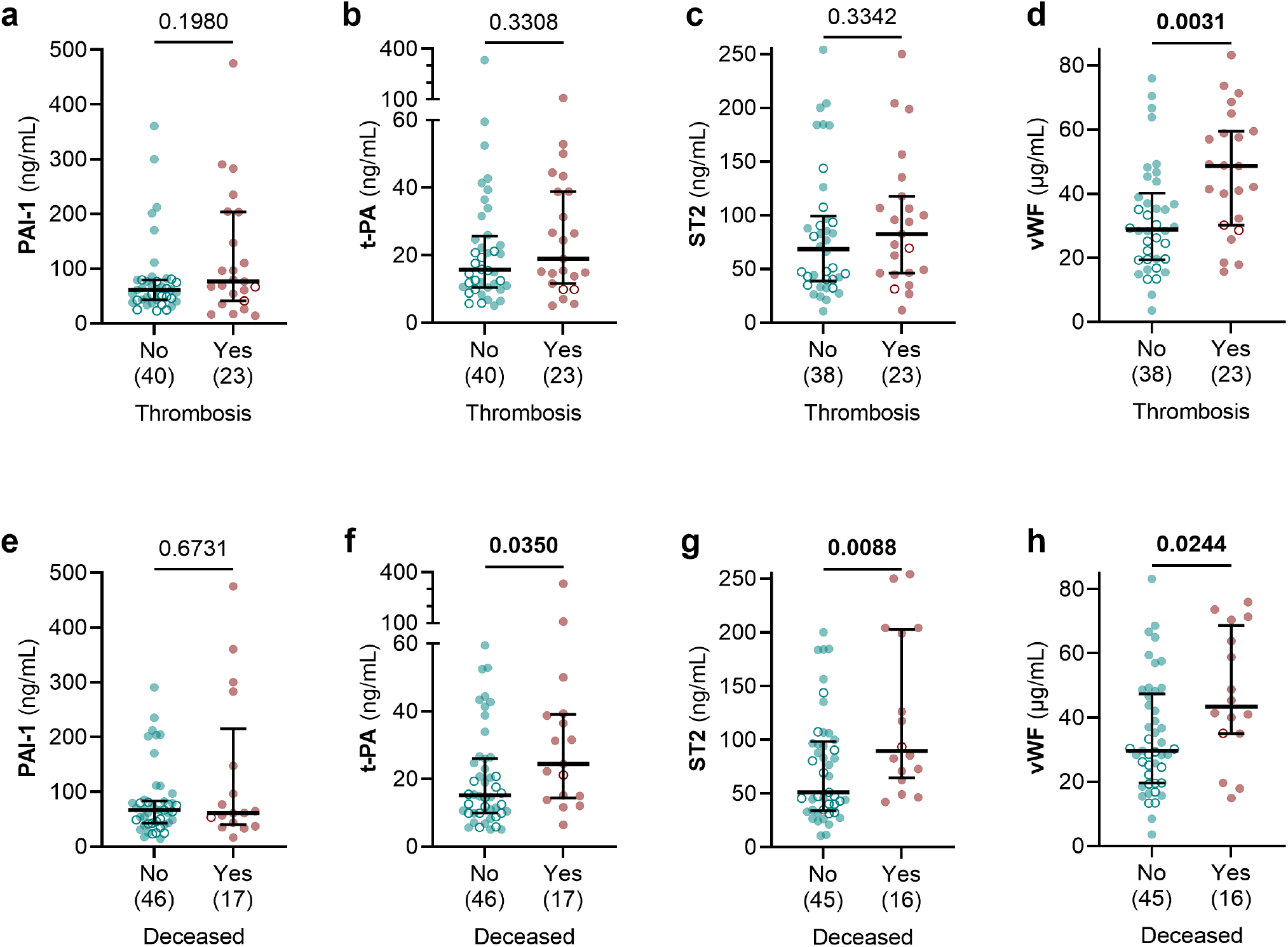
Higher levels of t-PA, ST2, and vWF are associated with worse clinical outcomes in COVID-19 patients. Levels of PAI-1 (**a, e**), t-PA (**b, f**), ST2 (**c, g**), and vWF (**d, h**) were compared between COVID-19 patients grouped by the detection of thrombotic events (**a-d**) and by survival outcome (**e-h**). The number of patients is indicated in parenthesis under each graph. Graphs show individual values with the median and 95% confidence intervals. Open circles represent non-ICU patients, and filled circles represent ICU patients. Groups were compared using the Mann-Whitney test and P-values are shown in each panel. Significant differences (P < 0.05) are in bold.

## 4. Discussion

Our study demonstrated that ICU patients with COVID-19 have elevated levels of PAI-1, t-PA, and fibrinogen, as well as vWF and the IL-33 receptor, ST2, compared to controls; these findings can explain the frequent observation of fibrin-mediated hypercoagulability and thrombosis in severe COVID-19.

There is normally a homeostatic equilibrium between coagulation and fibrinolysis. Excessive fibrinolysis can result in abnormal bleeding, whereas a deficit in fibrinolysis can result in plaque formation, disseminated intravascular coagulopathy, stroke, and thrombosis [14]. Coagulopathy in COVID-19 has a distinctive pathophysiology that appears to include not only impaired fibrinolysis but also platelet aggregation, inflammation, and microthrombi [15] (**Figure 4**). Evaluation of additional biomarkers studied here such as ST2 and vWF may help to characterize the COVID-19 associated coagulopathy and to identify patients at risk for worse clinical outcomes.

**Figure 4.**
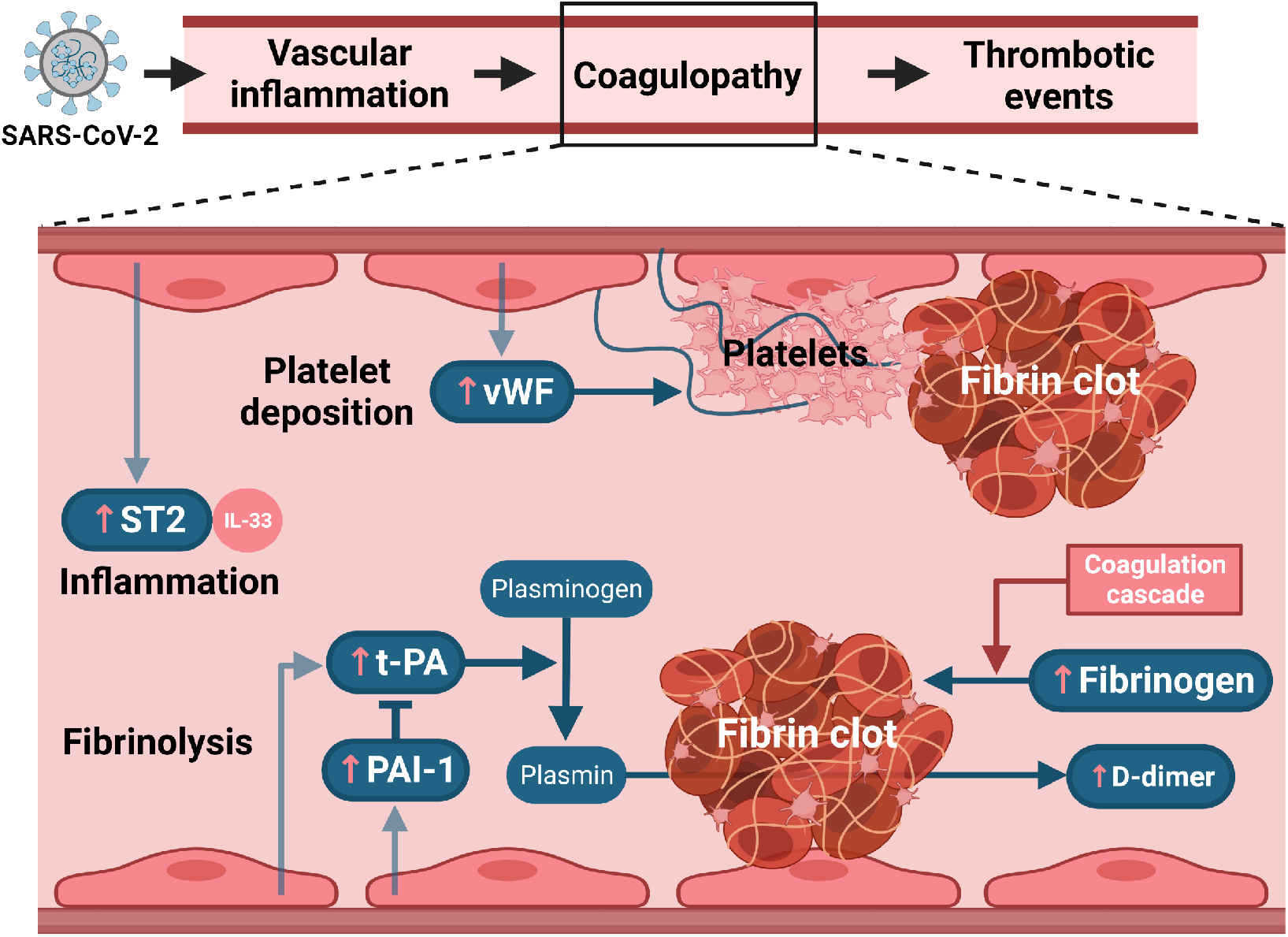
Role of coagulopathy markers in clot formation after SARS-CoV-2 infection. Diagram of key components of fibrinolysis, inflammation, and platelet deposition measured in this study. Pink up arrows depict changes of biomarkers in plasma of COVID-19 patients. Created with BioRender.com.

We measured elevated levels of fibrinogen and D-dimer in COVID-19 patients, as reported in similar studies [10,16], and these results are completely consistent with our prior observations on the important contribution of fibrinogen to the significant increase in clot viscosity in severe COVID-19 [8]. Increased production of fibrinogen, but not plasminogen, may also reflect the increase of extra-hepatically synthesis of fibrinogen in response to inflammation [17]. Together with elevated levels of fibrinogen and D-dimer, a “fibrinolysis shutdown” might also be a characteristic of severe cases of COVID-19, as proposed by previous authors [18].

One of the most important inhibitors of this fibrinolytic system is PAI-1 [19], and elevated levels of PAI-1 have been shown to increase the risk of atherothrombotic events [20]. Our observation of increased PAI-1 levels in COVID-19 further supports the hypothesis that reduced fibrinolysis caused by underproduction of plasmin may contribute to the hypercoagulability in COVID-19. In agreement with others [16,21], we also found an increase in t-PA levels, the enzyme responsible for the conversion of plasminogen into plasmin, in COVID-19 patients. This may lead to an increase in the rare but serious cases of bleeding in some patients [21] and very high t-PA levels were associated with non-survival in our study.

PAI-1, like t-PA, is normally produced and released from the vascular endothelium, but PAI-1 can also be produced by extravascular tissues, where its expression can be induced via activation of Nuclear Factor κB (NF-κB) by IL-6, Tumor necrosis factor α (TNF α) and a wide range of other pro-inflammatory mediators [19]. These mediators are known as components of the proposed cytokine storm syndrome that is the characteristic of many patients with severe COVID-19 [2].

The IL-33 acts on its cognate receptor ST2 and the IL-33/ST2 axis has been suggested to play a key role in the cytokine storm reported in COVID-19 [11]. We found high concentrations of ST2 in the plasma of COVID-19 patients, which has been suggested to be related to endothelial or pneumocyte inflammation and damage [11]. We found that higher levels of ST2 were associated with mortality, which supports the prognostic value of this biomarker in COVID-19, as suggested elsewhere [22].

Mast cells can play a role in the recruitment of inflammatory cells during vascular injury [23] and the production of PAI-1 [24]. Tryptase is generally measured as a proxy indicator of mast cell degranulation [12]. We did not find increases in tryptase in COVID-19 patients, but the involvement of mast cell degranulation in early stages of COVID-19 cannot be completely discarded, since tryptase has a short half-life of 2 hours and may trigger other factors involved in degranulation [12]. Nevertheless, tryptase does not seem likely to be a useful biomarker.

It has been suggested that the association between inflammation and thrombosis in COVID-19 might be mediated by endothelium injury and platelet aggregation [13]. The high levels of vWF found in the plasma of COVID-19 patients confirm similar results from recent studies [25,26]. These results might reflect an alteration in the balance of vWF and the protease ADAMTS13 (A Disintegrin And Metalloprotease with ThromboSpondin 1 repeats, number 13) that cleaves the vWF multimers released after endothelial damage [26,27]. The increased level of vWF may lead to platelet aggregation and a higher risk of microthrombi [13,27]. The association of high levels of vWF with thrombosis in our study would seem to support this hypothesis. Direct or indirect endothelial damage by SARS-CoV-2 and dysregulation of fibrinolysis have been suggested as additional mechanisms related to COVID-19 coagulopathy [7], and vWF may be a part of an endothelium-mediated inflammatory response, along with PAI-1, which may lead to worse clinical outcomes and survival rate, as seen in our study. In contrast, plasma levels of platelet activity marker PF4 and platelet count were not elevated in our cohort of patients, and this might be a diagnostic feature of COVID-19 that is distinct from sepsis-associated disseminated intravascular coagulation [28]. PF4 has drawn attention in relation to very rare cases of vaccine-induced thrombocytopenia, but this factor appears to be quite unrelated to the pathophysiology of COVID-19.

From the early stages of the COVID-19 pandemic, there have been attempts to rectify the complications associated with COVID-19 coagulopathy by stimulating fibrinolysis, with the initial focus exclusively on t-PA [29]. Recent results from a clinical trial of alteplase showed no bleeding events associated with the treatment but only a slight improvement in oxygenation [30]. It seems unlikely that the use of t-PA would be useful in COVID-19 patients with elevated levels of t-PA, as shown here and by others [21], potentially exposing these patients to excessive bleeding risk. Determining levels of t-PA and PAI-1 and calculating the ratio between t-PA and PAI-1 may help to some degree in identifying patients at risk for bleeding. Furthermore, t-PA administered exogenously has a short half-life [31] and its administration in severe COVID-19 patients would likely result in rapid inactivation, due to the very high levels of ambient PAI-1. There is evidence that streptokinase administered via a nebulizer can be effective in treating coagulopathy in ARDS resulting from other diseases [32] and it might be considered as a useful alternative treatment for COVID-19 coagulopathy.

A primary excess of PAI-1 may contribute to an imbalance between coagulation and fibrinolysis, resulting in the widespread and persistent coagulopathy seen in some COVID-19 patients. Inhibitors of PAI-1 such as tiplaxtinin (PAI-039) [33] or the novel TM5484 [34] are being studied as an intervention in COVID-19, supplementing the use of anticoagulants and corticosteroids. Measuring PAI-1 and t-PA levels, in addition to close monitoring for bleeding complications, would be required to assure safe use of any of these drugs. Other treatments related to the biomarkers measured in this study include anti-ST2 [11] and anti-vWF antibodies [35], but, to the best of our knowledge, clinical trials in COVID-19 are not yet underway.

Our study has some limitations, such as the modest sample size and the use of a single time point for measurement of the biomarkers. In addition, we suspect that the incidence of thrombosis might be underestimated due to the difficulties in the diagnosis of certain thrombotic events such as microthrombosis [36]. There have been some changes in medication regimens given to patients between the initial and subsequent waves of COVID-19. These might have some effect in the plasma levels of the biomarkers measured in this study and will be the subject of a later report. Nevertheless, the reproducible nature of our findings, and those of others, especially on PAI-1 [16,21] and vWF [26,27] supports the idea that high plasma levels of these biomarkers are a consistent feature of severe COVID-19.

Our study provides strong evidence that elevated levels of certain biomarkers such as t-PA, ST2, and vWF, as well as abnormal coagulation [8], may be related to the incidence of thrombosis, renal failure, and might contribute to other longer-term complications in COVID-19 patients. These biomarkers show robust and consistent increases in COVID-19 and can be assessed at the time of admission to non-ICU and ICU settings; they may prove useful as additional predictors of those patients at elevated risk for thrombosis or severe clinical outcomes. In a subsequent report, we will describe effects on these biomarkers and clinical outcomes of a number of medications used in COVID19.

## Data Availability

All data are available from the corresponding authors upon reasonable request

## Data Availability

All data are available from the corresponding authors upon reasonable request.

## Author contributions

NLH, GW, PY and DCG conceived the study and designed the experiments. AM, KE and GW recruited and consented patients. SP, KE, PY and AM retrieved patient data. GW, KE, DCG, NLH and MM processed samples, and DCG performed the ELISA experiments and data analysis. All authors contributed to data analysis and interpretation. NLH, DCG and GW wrote the first draft of the manuscript and all authors commented on and edited the manuscript.

## Acknowledgements

This work was supported solely by the Department of Anesthesiology at Columbia University Irving Medical Center (CUIMC). We are grateful to M. Shelanski and A. Brambrink for logistical support of our laboratories during the pandemic, and to C.W. Emala for many helpful discussions. We are deeply grateful to the patients and their families for their participation in this study.

## Supplementary figure

**Figure S1.**
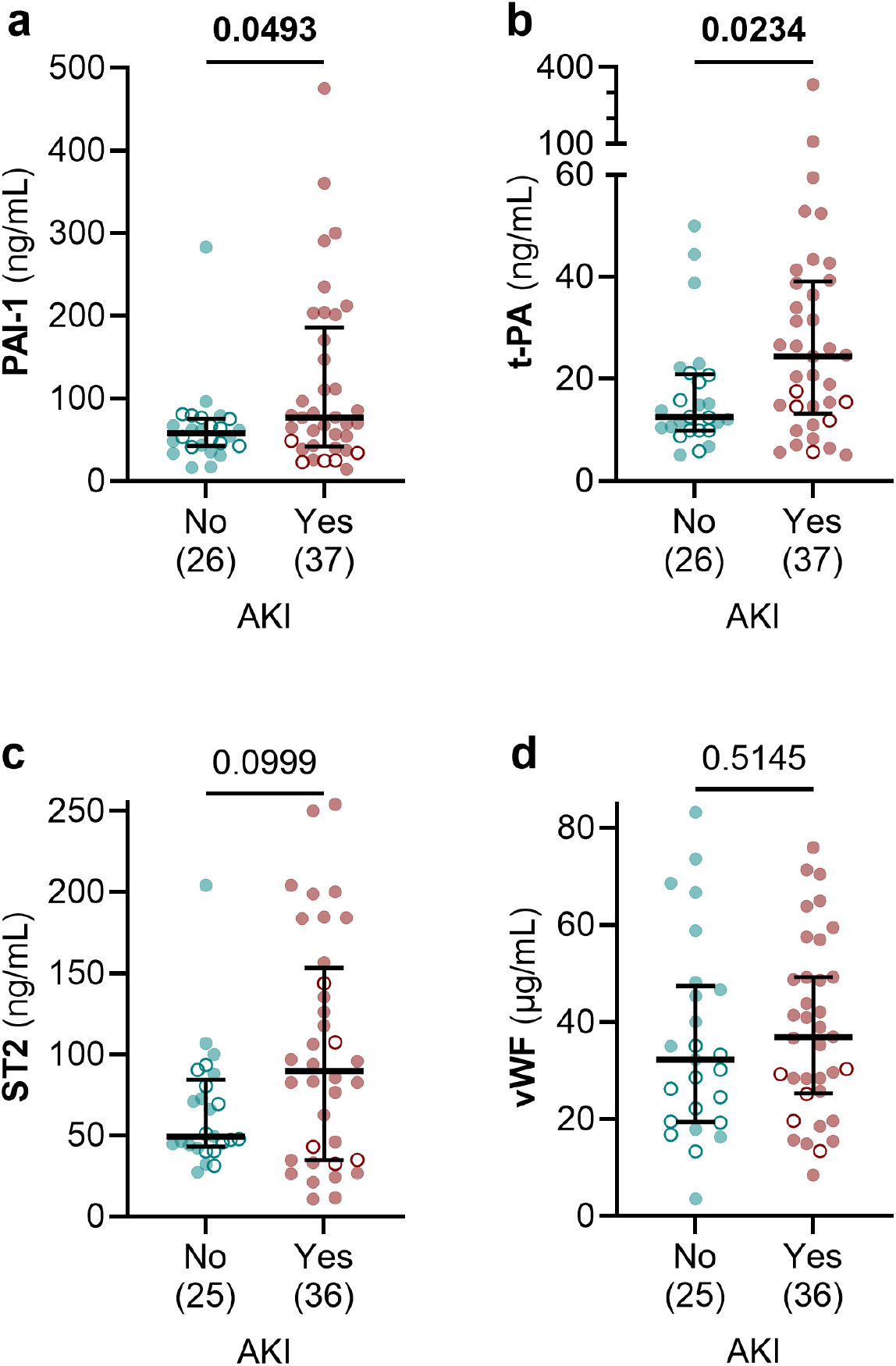
Higher levels of PAI-1 and t-PA were associated with AKI in COVID-19 patients. Levels of PAI-1 (**a**), t-PA (**b**), ST2 (**c**), ST2 (**e**), and vWF (**d**) were compared between COVID-19 patients grouped by the diagnosis of AKI. The number of patients is indicated in parenthesis under each graph. Graphs show individual values with the median and 95% confidence intervals. Open circles represent non-ICU patients, and filled circles represent ICU patients. Groups were compared using the Mann-Whitney test and P-values are shown in each panel. Significant differences (P < 0.05) are in bold.

